# Ethnicity-Specific Molecular Alterations in MAPK and JAK/STAT Pathways in Early-Onset Colorectal Cancer

**DOI:** 10.1101/2025.02.17.25322443

**Authors:** Cecilia Monge, Brigette Waldrup, Francisco G. Carranza, Enrique Velazquez-Villarreal

## Abstract

**Background/Objectives:** Early-onset colorectal cancer (EOCRC), defined as colorectal cancer (CRC) diagnosed before the age of 50, has been increasing in incidence, particularly among Hispanic/Latino (H/L) populations. Despite this trend, the underlying molecular mechanisms driving EOCRC disparities remain poorly understood. The MAPK and JAK/STAT pathways play critical roles in tumor progression, proliferation, and treatment response; however, their involvement in ethnicity-specific differences in EOCRC remains unclear. This study aims to characterize molecular alterations in MAPK and JAK/STAT pathway genes among EOCRC patients, focusing on differences between H/L and Non-Hispanic White (NHW) patients. Additionally, we assess whether these pathway-specific alterations contribute to survival outcomes in H/L EOCRC patients.

**Methods:** We conducted a bioinformatics analysis using publicly available CRC datasets to assess mutation frequencies in MAPK and JAK/STAT pathway genes. A total of 3,412 patients were included in the study, comprising 302 H/L patients and 3,110 NHW patients. Patients were stratified by age (EOCRC: <50 years, late-onset colorectal cancer –LOCRC: ≥50 years) and ethnicity (H/L vs. NHW) to evaluate differences in mutation prevalence. Chi-squared tests were performed to compare mutation rates between groups, and Kaplan-Meier survival analysis was used to assess overall survival differences based on pathway alterations among both H/L and NHW EOCRC patients.

**Results:** Significant differences were observed in MAPK pathway-related genes when comparing EOCRC and LOCRC in H/L patients. NF1 (11.6% vs. 3.7%, p = 0.01), ACVR1 (2.9% vs. 0%, p = 0.04), and MAP2K1 (3.6% vs. 0%, p = 0.01) were more prevalent in EOCRC, while BRAF mutations (18.3% vs. 5.1%, p = 9.1e-4) were significantly more frequent in LOCRC among H/L patients. Additionally, when comparing EOCRC in H/L patients to EOCRC in NHW patients, key MAPK pathway genes such as AKT1 (5.1% vs. 1.8%, p = 0.03), MAPK3 (3.6% vs. 0.7%, p = 6.83e-3), NF1 (11.6% vs. 6.1%, p = 0.02), and PDGFRB (5.8% vs. 2.1%, p = 0.02) were significantly enriched in H/L EOCRC patients.

However, no significant differences were observed in JAK/STAT pathway-related genes when comparing EOCRC and LOCRC in H/L patients, nor when comparing EOCRC in H/L vs. NHW patients. Survival analysis revealed borderline significant differences in H/L EOCRC patients, whereas NHW EOCRC patients with no alterations in the JAK/STAT pathway exhibited significant survival differences. In contrast, MAPK pathway alterations were not associated with significant survival differences. These findings suggest that MAPK and JAK/STAT pathway disruptions may have distinct prognostic implications in H/L EOCRC patients, justifying further investigation into their potential role in cancer progression and treatment response.

**Conclusions:** These findings suggest that MAPK pathway dysregulation plays a distinct role in EOCRC among H/L patients, potentially contributing to disparities in CRC development and treatment response. The higher prevalence of MAPK alterations in H/L EOCRC patients compared to NHW patients underscores the need to explore ethnic-specific tumor biology and therapeutic targets. Conversely, the lack of significant differences in JAK/STAT pathway alterations suggests that this pathway may not play a major differential role in EOCRC vs. LOCRC within this population. Survival analysis highlighted the prognostic relevance of pathway-specific disruptions. These insights emphasize the importance of precision medicine approaches that consider genetic heterogeneity and pathway-specific alterations to improve outcomes for H/L CRC patients.

## 1. Introduction

Colorectal cancer (CRC) is the third most common cancer worldwide and the second leading cause of cancer-related mortality (1, 2). While the overall incidence of CRC has stabilized or declined in higher-income countries, an alarming rise in early-onset colorectal cancer (EOCRC), defined as CRC diagnosed before the age of 50, has been observed globally (1,3–5). This trend is particularly pronounced in the Hispanic/Latino (H/L) population, which has experienced the most significant increase in EOCRC incidence among all racial and ethnic groups in the United States (6, 7). Furthermore, H/L individuals have the highest increase in EOCRC-related mortality compared to non-Hispanic White (NHW) populations, highlighting a critical health disparity (8,10). These disparities underscore the urgent need for a deeper understanding of the molecular mechanisms contributing to CRC development and progression in H/L patients.

Screening guidelines that recommend routine CRC surveillance beginning at age 50 may contribute to late-stage diagnoses in EOCRC patients (11). Beyond delayed detection, emerging research suggests that EOCRC has distinct molecular characteristics compared to late-onset colorectal cancer (LOCRC), including a higher prevalence of microsatellite instability (MSI), an increased tumor mutation burden, and elevated expression of immune checkpoint regulators such as PD-L1 (12–15). Additionally, epigenetic alterations, such as LINE-1 hypomethylation, have been proposed as biomarkers that distinguish EOCRC from LOCRC (16). These findings suggest that EOCRC may develop through distinct oncogenic pathways, justifying further investigation into ethnicity-specific genomic alterations.

The mitogen-activated protein kinase (MAPK) and Janus kinase/signal transducer and activator of transcription (JAK/STAT) pathways are two critical signaling cascades involved in CRC pathogenesis, influencing cell proliferation, differentiation, survival, and immune response regulation. The MAPK pathway comprises four primary signaling families—ERK, BMK-1, JNK, and p38—all of which regulate key oncogenic processes (16). In CRC, aberrant MAPK signaling is frequently driven by oncogenic mutations in RAS and BRAF, leading to uncontrolled tumor growth, resistance to apoptosis, and metastasis (17, 18). Notably, BRAF mutations, which are common in CRC, have been shown to confer resistance to RAF inhibitors, such as vemurafenib, due to EGFR-mediated reactivation of the MAPK pathway (19, 20). While the role of the MAPK pathway in CRC is well established, its contribution to EOCRC disparities in H/L patients remains poorly understood.

The JAK/STAT pathway is another essential signaling cascade that mediates cellular responses to inflammation, an established driver of CRC progression. Persistent activation of JAK/STAT signaling has been implicated in tumor progression, therapy resistance, and poor clinical outcomes in CRC patients (21). Elevated JAK/STAT activity has been associated with increased cytokine production, enhanced immune evasion, and epithelial-to-mesenchymal transition (EMT) in CRC, ultimately promoting tumor aggressiveness (21). Additionally, STAT3 activation in CRC has been linked to stromal invasion and reduced cancer-specific survival, particularly in tumors with high inflammatory signaling (22–24). Despite its significance, the role of JAK/STAT pathway alterations in EOCRC, particularly among H/L patients, remains unexplored.

Given the increasing burden of EOCRC in the H/L population and the limited knowledge of ethnicity-specific oncogenic mechanisms, this study aims to characterize molecular alterations in the MAPK and JAK/STAT pathways in EOCRC. By comparing EOCRC in H/L and NHW patients, we seek to identify key pathway-specific differences that may contribute to CRC disparities. Additionally, we evaluate the impact of these alterations on patient survival, providing insights into potential prognostic biomarkers and therapeutic targets. Understanding the molecular landscape of EOCRC in underrepresented populations is essential for advancing precision medicine approaches and improving outcomes in H/L CRC patients.

## 2. Materials and Methods

This study utilized clinical and genomic data from 20 CRC datasets available through the cBioPortal database. The datasets analyzed included colorectal adenocarcinoma, colon adenocarcinoma, and rectal adenocarcinoma, as well as data from the GENIE BPC CRC v2.0-public dataset. To ensure a focus on primary tumor cases, two datasets that specifically examined metastatic CRC were excluded. Patients were selected based on predefined inclusion criteria, which required identification as Hispanic or Latino, Spanish, NOS; Hispanic, NOS; Latino, NOS; or individuals with a Mexican or Spanish surname. Additional filtering criteria included limiting cases to primary tumors, selecting colorectal, colon, and rectal adenocarcinomas, confirming adenocarcinoma, NOS histology, and ensuring only one sample per patient. Three datasets met all these selection criteria: TCGA PanCancer Atlas, MSK Nat Commun 2022, and GENIE BPC CRC, yielding a cohort of 302 H/L patients (138 EOCRC and 164 LOCRC). Similarly, 3,110 NHW patients (897 EOCRC and 2,213 LOCRC) were identified using the same inclusion criteria (Tables 1 & 2). Age at diagnosis was obtained from GENIE database clinical records. This study represents one of the largest investigations into MAPK and JAK/STAT pathway alterations in an underserved population, providing essential insights into the molecular disparities between EOCRC and LOCRC patients.

**Table 1.** Demographic and clinical characteristics of Hispanic/Latino (H/L) and Non-Hispanic White (NHW) cohorts.

| Clinical Feature | H/L Cohort<br>n (%) | NHW Cohort<br>n (%) |
| --- | --- | --- |
| **Age Onset and Gender** |  |  |
| Early-Onset (< 50) Male | 83 (27.5%) | 503 (16.2%) |
| Early-Onset (< 50) Female | 55 (18.2%) | 394 (12.7%) |
| Late-Onset (≥ 50) Male | 93 (30.8%) | 1209 (38.9%) |
| Late-Onset (≥ 50) Female | 71 (23.5%) | 1004 (32.3%) |
| **Stage at Diagnosis** |  |  |
| Stage 1-3 | 98 (32.5%) | 965 (31.0%) |
| Stage 4 | 132 (43.7%) | 1131 (36.4%) |
| NA | 72 (23.8%) | 1014 (32.6%) |
| **Ethnicity** |  |  |
| Spanish NOS; Hispanic NOS, Latino NOS | 270 (89.4%) | 0 (0.0%) |
| Mexican (includes Chicano) | 28 (9.3%) | 0 (0.0%) |
| Other Spanish/Hispanic | 1 (0.3%) | 0 (0.0%) |
| Spanish surname only | 3 (1.0%) | 0 (0.0%) |
| Non-Spanish; Non-Hispanic | 0 (0.0%) | 3110 (100.0%) |

**Table 2.** Ethnicity-associated differences in clinical characteristics between Hispanic/Latino (H/L) and Non-Hispanic White (NHW) cohorts.

| Clinical Feature | Early-Onset HL<br>n (%) | Late-Onset HL<br>n (%) | p-value | Early-Onset HL<br>n (%) | Early-Onset NHW<br>n (%) | p-value |
| --- | --- | --- | --- | --- | --- | --- |
| Median Diagnosis Age (IQR) | 41 (36-46) | 61 (55-69) | < **0.05** | 41 (36-46) | 42 (38-47) | < **0.05** |
| Median Mutation Count* | 7 (5-10) | 8 (6-10) | > 0.05 | 7 (5-10) | 7 (5-10) | > 0.05 |
| ACVR1 Mutation |  |  |  |  |  |  |
| Present | 4 (2.9%) | 0 (0.0%) | < **0.05** | 4 (2.9%) | 11 (1.2%) | > 0.05 |
| Absent | 134 (97.1%) | 164 (100.0%) |  | 134 (97.1%) | 886 (98.8%) |  |
| AKT1 Mutation |  |  |  |  |  |  |
| Present | 7 (5.1%) | 3 (1.8%) | > 0.05 | 7 (5.1%) | 16 (1.8%) | < **0.05** |
| Absent | 131 (94.9%) | 161 (98.2%) |  | 131 (94.9%) | 881 (98.2%) |  |
| BRAF Mutation |  |  |  |  |  |  |
| Present | 7 (5.1%) | 30 (18.3%) | < **0.05** | 7 (5.1%) | 67 (7.5%) | > 0.05 |
| Absent | 131 (94.9%) | 134 (81.7%) |  | 131 (94.9%) | 830 (92.5%) |  |
| MAPK2K1 Mutation |  |  |  |  |  |  |
| Present | 5 (3.6%) | 0 (0.0%) | < **0.05** | 5 (3.6%) | 16 (1.8%) | > 0.05 |
| Absent | 133 (96.4%) | 164 (100.0%) |  | 133 (96.4%) | 881 (98.2%) |  |
| MAPK3 Mutation |  |  |  |  |  |  |
| Present | 5 (3.6%) | 1 (0.6%) | > 0.05 | 5 (3.6%) | 6 (0.7%) | < **0.05** |
| Absent | 133 (96.4%) | 163 (99.4%) |  | 133 (96.4%) | 891 (99.3%) |  |
| NF1 Mutation |  |  |  |  |  |  |
| Present | 16 (11.6%) | 6 (3.7%) | < **0.05** | 16 (11.6%) | 55 (6.1%) | < **0.05** |
| Absent | 122 (88.4%) | 158 (96.3%) |  | 122 (88.4%) | 842 (93.9%) |  |
| PDGFRB Mutation |  |  |  |  |  |  |
| Present | 8 (5.8%) | 3 (1.8%) | > 0.05 | 8 (5.8%) | 19 (2.1%) | < **0.05** |
| Absent | 130 (94.2%) | 161 (98.2%) |  | 130 (94.2%) | 878 (97.9%) |  |

\*LO HL: NA 1, EO NHW: NA 8

Pathway alterations were defined based on previously established criteria (18). Patients were categorized into EOCRC (<50 years of age) and LOCRC (≥50 years) groups, with further stratification based on ethnicity (H/L vs. NHW) and pathway status (presence or absence of MAPK and JAK/STAT pathway alterations). Table 3 presents the distribution of EOCRC and LOCRC H/L CRC patients, highlighting the prevalence of MAPK and JAK/STAT pathway alterations in each group. Table 4 extends this analysis by comparing EOCRC H/L and EOCRC NHW patients, facilitating a comparative assessment of pathway-specific differences between ethnic groups. Through these stratifications, this study offers a comprehensive molecular characterization of MAPK and JAK/STAT pathway alterations, providing valuable insights into potential ethnic-specific disparities that could inform precision medicine strategies for CRC.

**Table 3.** Frequency of MAPK and JAK/STAT pathway alterations in Early-Onset (EOCRC) and Late-Onset (LOCRC) Colorectal Cancer among Hispanic/Latino (H/L) patients.

|  | Early-Onset H/L<br>n (%) | Late-Onset H/L<br>n (%) | p-value |
| --- | --- | --- | --- |
| JAK/STAT Alterations Present | 16 (11.6%) | 19 (11.6%) | 1 |
| JAK/STAT Alterations Absent | 122 (88.4%) | 145 (88.4%) |  |
| MAPK Alterations Present | 134 (97.1%) | 161 (98.2%) | 0.7063 |
| MAPK Alterations Absent | 4 (2.9%) | 3 (1.8%) |  |

**Table 4.** Frequency of MAPK and JAK/STAT pathway alterations in Early-Onset Colorectal Cancer (EOCRC) among Hispanic/Latino (H/L) and Non-Hispanic White (NHW) patients.

|  | Early Onset H/L<br>n (%) | Early Onset NHW<br>n (%) | p-value |
| --- | --- | --- | --- |
| JAK/STAT Alterations Present | 16 (11.6%) | 78 (8.7%) | 0.3452 |
| JAK/STAT Alterations Absent | 122 (88.4%) | 819 (91.3%) |  |
| MAPK Alterations Present | 137 (99.3%) | 861 (96.0%) | 0.6604 |
| MAPK Alterations Absent | 4 (2.9%) | 36 (4.0%) |  |

Statistical comparisons of mutation frequencies between groups were conducted using Chi-square tests to assess the independence of categorical variables and evaluate associations between age, ethnicity, and pathway alterations. Additionally, tumor samples were further stratified based on tumor location (colon vs. rectal adenocarcinoma), allowing for a more refined analysis of the interplay between tumor site, ethnicity, and pathway alterations. This level of stratification enabled a nuanced investigation into patient heterogeneity and potential implications for treatment responses.

To assess the prognostic relevance of MAPK and JAK/STAT pathway alterations, Kaplan-Meier survival analysis was performed to evaluate overall survival across different patient subgroups. Survival curves were constructed to illustrate differences in survival probability over time, with patients grouped based on the presence or absence of pathway alterations. The log-rank test was applied to determine statistically significant differences between survival curves, and median survival times were calculated alongside 95% confidence intervals (CIs) to estimate survival precision. By integrating large-scale genomic data, survival analysis, and stratified comparisons, this study provides a detailed assessment of pathway-specific disruptions in EOCRC and LOCRC patients, particularly within the H/L population, offering new insights into ethnicity-specific molecular mechanisms in CRC.

## 3. Results

From three cBioPortal projects that reported ethnicity, we constructed our H/L cohort, which comprised 302 samples, while the NHW cohort included 3,110 samples. In the H/L cohort, 27.5% of patients were diagnosed with EOCRC (before age 50), while 72.5% were diagnosed at age 50 or older. In contrast, the NHW cohort had 16.2% EOCRC patients, with the majority (83.8%) diagnosed at age 50 or older (Table 1). The gender distribution within the H/L cohort included 58.3% male and 41.7% female patients, whereas the NHW cohort consisted of 55% male and 45% female patients. At the time of diagnosis, 32.5% of patients in the H/L cohort were diagnosed at stages 0, I, II, or III, while 43.7% had stage IV disease. In comparison, the NHW cohort had 31% of patients diagnosed at stages 0, I, II, or III, and 36.4% at stage IV. Notably, 23.8% of H/L patients and 32.6% of NHW patients were recorded as NA for stage at diagnosis, indicating missing or unreported staging information. Ethnicity classification within the H/L cohort revealed that the majority of patients (89.4%) identified as Spanish NOS; Hispanic NOS; or Latino NOS, while 9.3% were classified as Mexican (including Chicano), and 1.3% identified as either Other Spanish/Hispanic or Spanish surname only. Conversely, all patients in the NHW cohort (100%) were classified as NHW, ensuring a clear distinction between the two groups for comparative analyses.

A comparison of clinical features between EOCRC and LOCRC H/L patients, as well as between EOCRC H/L and EOCRC NHW patients, identified significant distinctions (Table 2). The median age at diagnosis for EOCRC H/L patients was 41 years (IQR: 36– 46), significantly younger than the median of 61 years (IQR: 55–69) observed in LOCRC H/L patients (p<0.05). Similarly, EOCRC H/L patients were diagnosed at a significantly younger age compared to EOCRC NHW patients (median: 42 years, IQR: 38–47) (p<0.05). Analysis of mutation burden revealed a median mutation count of 7 in EOCRC H/L patients, compared to a slightly higher median count of 8 in LOCRC H/L patients; however, this difference was not statistically significant (p>0.05). Similarly, EOCRC NHW patients had a median mutation count of 7, comparable to EOCRC H/L patients, with no statistically significant difference between these groups (p>0.05).

Regarding pathway-specific alterations, several genes within the MAPK pathway were found to be more frequently mutated in EOCRC H/L patients than in their LOCRC counterparts. Notably, NF1 (11.6% vs. 3.7%, p = 0.01), ACVR1 (2.9% vs. 0%, p = 0.04), and MAP2K1 (3.6% vs. 0%, p = 0.01) exhibited significantly higher mutation frequencies in EOCRC. In contrast, BRAF mutations were significantly more common in LOCRC H/L patients compared to EOCRC patients (18.3% vs. 5.1%, p = 9.1e-4). When comparing EOCRC in H/L versus NHW patients, key MAPK pathway genes such as AKT1 (5.1% vs. 1.8%, p = 0.03), MAPK3 (3.6% vs. 0.7%, p = 6.83e-3), NF1 (11.6% vs. 6.1%, p = 0.02), and PDGFRB (5.8% vs. 2.1%, p = 0.02) were significantly enriched in EOCRC H/L patients. These findings underscore potential ethnic-specific genomic differences that may contribute to variations in CRC pathogenesis.

The observed disparities in MAPK pathway alterations between EOCRC and LOCRC, as well as between H/L and NHW EOCRC patients, highlight the need for further investigation into the biological and clinical implications of these mutations. Understanding these differences may help inform targeted therapeutic strategies aimed at addressing disparities in CRC outcomes across diverse ethnic populations.

In our analysis of genetic alterations among H/L individuals with EOCRC and LOCRC, no significant differences were observed in the frequency of MAPK and JAK/STAT pathway alterations (Table 3). JAK/STAT pathway alterations were present in 11.6% of both EOCRC and LOCRC patients, with an identical absence rate of 88.4% in both groups. Similarly, MAPK pathway alterations were highly prevalent in both groups, occurring in 97.1% of early-onset patients and 98.2% of late-onset patients, though this difference was not statistically significant (p = 0.7). The absence of MAPK pathway alterations was slightly more common in EORCR patients (2.9%) compared to LOCRC patients (1.8%). These findings suggest that MAPK and JAK/STAT pathway mutations are consistently frequent across both age groups, indicating that their role in CRC development may not be significantly influenced by age at onset in H/L patients. Further studies are justified to explore potential interactions between these pathways and other molecular drivers contributing to CRC progression in this population.

In our analysis of genetic alterations in EOCRC among H/L and NHW individuals, no statistically significant differences were observed in the frequency of MAPK and JAK/STAT pathway mutations. JAK/STAT alterations were detected in 11.6% of EOCRC H/L patients compared to 8.7% of EOCRC NHW patients (p = 0.34). The absence of JAK/STAT alterations was slightly more common in NHW individuals (91.3%) than in H/L individuals (88.4%), though this difference was not statistically significant (p = 0.34).

Similarly, MAPK pathway alterations were highly prevalent in both EOCRC groups, occurring in 99.3% of H/L patients and 96% of NHW patients (p = 0.66). The absence of MAPK pathway alterations was slightly more frequent in NHW individuals (4%) than in H/L individuals (2.9%), but this difference also lacked statistical significance (p = 0.34). These findings suggest that MAPK and JAK/STAT pathway mutations may be marginally more frequent in EOCRC H/L individuals than in their NHW counterparts, but the observed differences are not statistically significant. Further analyses incorporating larger cohorts and functional studies are needed to better understand the clinical relevance of these pathway alterations and their potential implications for precision medicine and targeted therapies in CRC.

The Kaplan-Meier survival analysis for EOCRC H/L CRC patients revealed no statistically significant differences in overall survival between those with and without MAPK pathway alterations (Figure 2). While the survival curves for patients with and without MAPK alterations appeared to diverge slightly over time, the p-value (p = 0.9) indicated that this difference was not statistically significant. The confidence intervals surrounding each curve reflect variability in survival estimates at different time points, underscoring the uncertainty of these findings. These results suggest that MAPK pathway alterations may not play a major role in influencing overall survival outcomes in EOCRC H/L patients. Further studies with larger sample sizes and additional molecular stratifications are needed to better understand the potential impact of MAPK pathway disruptions in this population.

**Figure 1.**
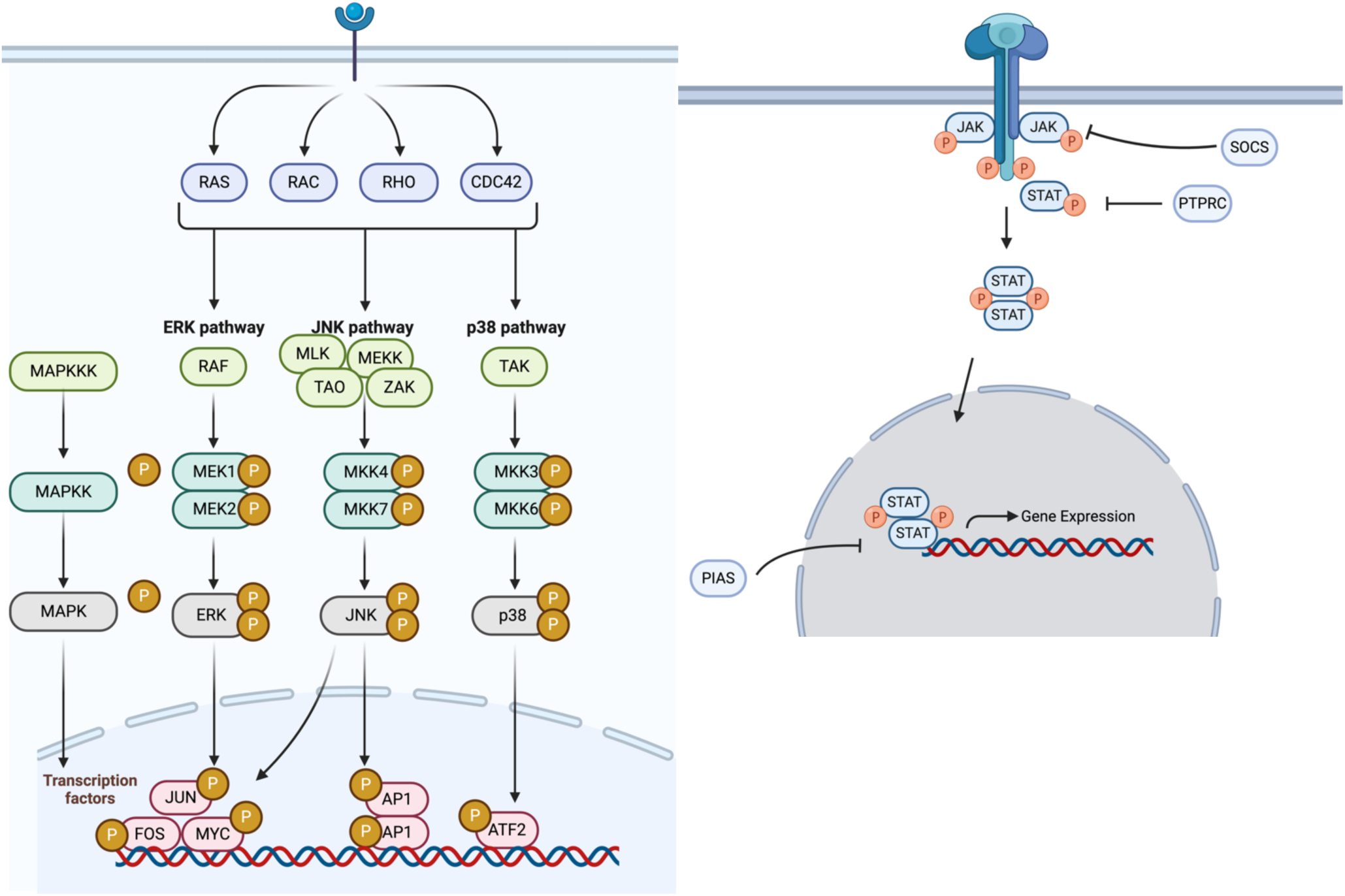
Illustration of the MAPK (left) and JAK/STAT (right) signaling pathways. The left panel depicts the MAPK (mitogen-activated protein kinase) signaling pathway, which plays a crucial role in cell proliferation, differentiation, survival, and stress response. This pathway is activated through various receptor-mediated signals, including RAS, RAC, RHO, and CDC42, leading to three major downstream cascades: ERK, JNK, and p38 pathways. The ERK pathway is primarily involved in cell proliferation and differentiation, whereas the JNK and p38 pathways regulate stress responses, apoptosis, and inflammation. Activation of these pathways results in the phosphorylation and activation of transcription factors such as JUN, FOS, MYC, AP1, and ATF2, which influence gene expression and tumor progression. Dysregulation of MAPK pathway genes, including NF1, BRAF, AKT1, and MAPK3, has been implicated in early-onset colorectal cancer (EOCRC), particularly among H/L patients. The right panel illustrates the JAK/STAT (Janus kinase/signal transducer and activator of transcription) pathway, a key intracellular signaling cascade involved in immune regulation, inflammation, and tumorigenesis. Ligand binding to cytokine receptors induces the phosphorylation of JAK kinases, which subsequently activate STAT proteins through phosphorylation. Phosphorylated STAT dimers translocate to the nucleus, where they regulate gene expression involved in cell proliferation, survival, and immune responses. This pathway is negatively regulated by SOCS and PTPN6, which act as feedback inhibitors.

**Figure 2.**
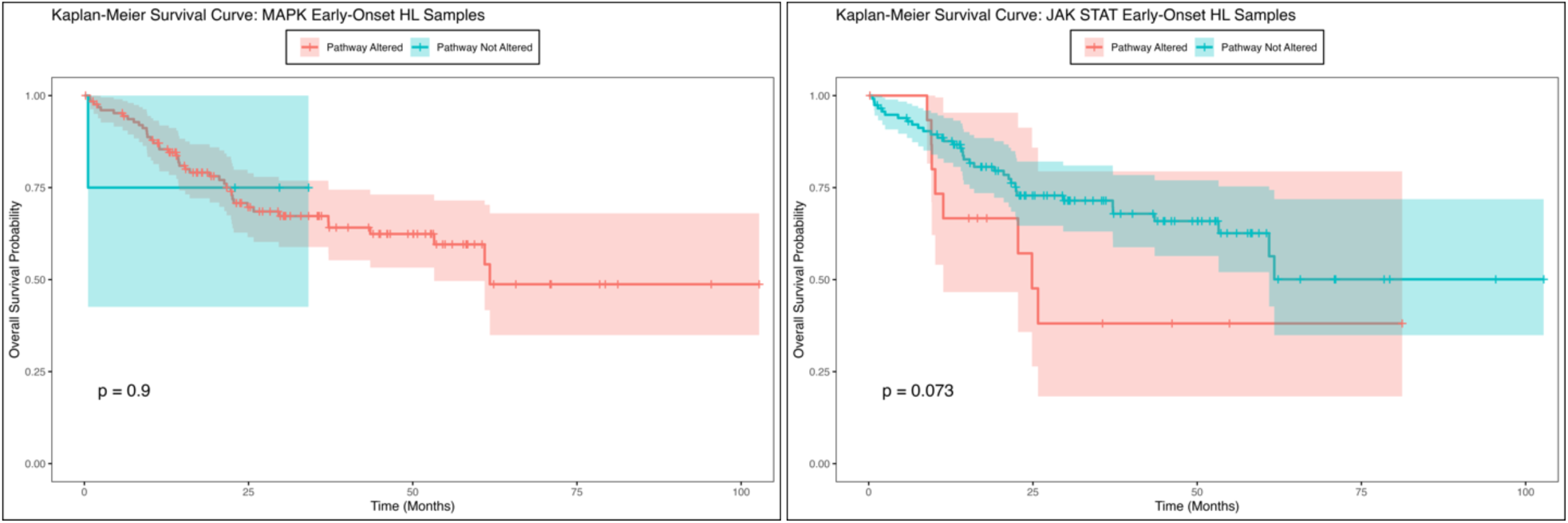
Overall survival curves of early-onset colorectal cancer (EOCRC) Hispanic/Latino (H/L) patients stratified by the presence or absence of MAPK (left) and JAK/STAT (right) pathway alterations. The left panel illustrates the Kaplan-Meier survival curve for EOCRC H/L patients stratified by MAPK pathway alterations. Patients with MAPK pathway alterations (red curve) exhibited no significant survival difference compared to those without alterations (blue curve) (p = 0.9). The shaded regions around the curves represent 95% confidence intervals, and vertical tick marks indicate censored patients. These results suggest that MAPK pathway disruptions may not have a strong prognostic impact on overall survival in EOCRC H/L patients. The right panel presents the Kaplan-Meier survival curve for EOCRC H/L patients stratified by JAK/STAT pathway alterations. Patients with JAK/STAT pathway alterations (red curve) showed a trend toward worse survival outcomes compared to those without alterations (blue curve), though this difference was only borderline significant (p = 0.073).

The Kaplan-Meier survival analysis for EOCRC H/L CRC patients with and without JAK/STAT pathway alterations (Figure 2) revealed a trend toward worse survival outcomes in patients with JAK/STAT alterations. Although the difference in overall survival did not reach statistical significance, the p-value (p = 0.073) suggests a borderline association. Patients with JAK/STAT pathway alterations exhibited an early decline in survival probability, with consistently lower survival rates throughout the follow-up period compared to those without alterations. In contrast, patients without JAK/STAT alterations demonstrated a more gradual decline in survival probability, maintaining higher overall survival rates over time. These findings indicate a potential prognostic impact of JAK/STAT pathway disruptions in EOCRC H/L patients, justifying further investigation. Larger cohorts and additional molecular characterizations are necessary to clarify the role of JAK/STAT alterations in survival outcomes within this population.

Similar to the results observed for JAK/STAT pathway in the H/L cohort, the overall survival analysis for the NHW cohort (Figure S1) suggests that JAK/STAT pathway alterations significantly impact survival outcomes in EOCRC within this ethnic group. The highly significant p-value (< 0.0001) indicates a strong association between JAK/STAT pathway disruptions and poorer survival, highlighting its potential prognostic relevance. In contrast, MAPK pathway alterations in NHW patients did not show a statistically significant difference in overall survival, with a p-value of 0.49. These findings contrast with those observed in the H/L cohort, where neither MAPK nor JAK/STAT alterations reached statistical significance for survival differences. This suggests that JAK/STAT pathway disruptions may play a more prominent role in EOCRC survival among NHW patients, emphasizing the need for further investigation into ethnicity-specific molecular drivers of CRC prognosis.

The alteration rates of MAPK and JAK/STAT pathway-related genes were analyzed among EOCRC and LOCRC H/L patients to determine potential age-related differences (Table S1). The analysis revealed that specific MAPK pathway genes, including NF1, ACVR1, and MAP2K1, were significantly more prevalent in EOCRC patients compared to LOCRC patients. NF1 alterations were observed in 11.6% of EOCRC patients, compared to only 3.7% in LOCRC patients (p = 0.01). Similarly, ACVR1 and MAP2K1 alterations were exclusive to EOCRC patients (2.9% and 3.6%, respectively) and were absent in LOCRC patients (p = 0.04 and p = 0.01, respectively). Conversely, BRAF mutations were significantly more frequent in LOCRC patients (18.3%) compared to EOCRC patients (5.1%, p = 9.1e-4), indicating a potential age-associated pattern in MAPK pathway gene alterations.

Comparing EOCRC H/L patients to their NHW counterparts, several key MAPK pathway genes exhibited significantly higher alteration rates in H/L patients (Table S3). AKT1 mutations were more frequent in EOCRC H/L patients (5.1%) compared to EOCRC NHW patients (1.8%, p = 0.03). Similarly, MAPK3 (3.6% vs. 0.7%, p = 6.83e-3), NF1 (11.6% vs. 6.1%, p = 0.02), and PDGFRB (5.8% vs. 2.1%, p = 0.02) showed a significantly higher prevalence in EOCRC H/L patients. In contrast, JAK/STAT pathway alterations did not show significant differences between EOCRC and LOCRC H/L patients or when comparing EOCRC H/L and NHW patients. These findings suggest that while alterations in the MAPK pathway may play a distinct role in EOCRC CRC among H/L patients, JAK/STAT pathway alterations appear to be less associated with age or ethnicity-specific differences. The significantly higher frequency of NF1, ACVR1, MAP2K1, and AKT1 mutations in EOCRC H/L patients underscores the need for further investigation into their functional roles in tumor progression and response to therapy.

Future studies should explore the implications of these molecular differences in precision medicine strategies tailored for H/L CRC patients across different age groups.

## 4. Discussion

The MAPK and JAK/STAT signaling pathways are fundamental regulators of cellular proliferation, differentiation, and survival, playing pivotal roles in CRC pathogenesis (18, 19). The MAPK pathway, frequently activated in CRC, drives tumorigenesis by promoting cell proliferation and resistance to apoptosis, while the JAK/STAT pathway is essential in mediating immune signaling and inflammatory responses in tumor progression (21, 22). Although these pathways have been extensively studied in CRC, their specific roles and ethnic variations in EOCRC remain underexplored, particularly among H/L patients.

Our study provides a comprehensive analysis of MAPK and JAK/STAT pathway alterations in EOCRC, highlighting significant differences in mutation frequencies between H/L and NHW patients. Notably, key MAPK pathway genes such as NF1 (11.6% vs. 6.1%, p = 0.02), MAPK3 (3.6% vs. 0.7%, p = 6.83e-3), AKT1 (5.1% vs. 1.8%, p = 0.03), and PDGFRB (5.8% vs. 2.1%, p = 0.02) were more frequently altered in H/L EOCRC patients compared to their NHW counterparts. These findings suggest that ethnic-specific genetic variations in MAPK signaling may contribute to CRC disparities, potentially influencing tumor progression and response to targeted therapies (18–20).

Interestingly, when comparing EOCRC and LOCRC within the H/L cohort, significant differences were observed in MAPK-related gene alterations. NF1, ACVR1, and MAP2K1 mutations were significantly more prevalent in EOCRC, while BRAF mutations were markedly higher in LOCRC patients. These findings suggest that MAPK dysregulation may play a unique role in EOCRC among H/L patients, possibly contributing to the aggressive nature of CRC in this population. However, JAK/STAT pathway alterations did not show significant differences between EOCRC and LOCRC, nor between H/L and NHW patients, indicating that this pathway (21, 22) may not be a major driver of ethnic disparities in CRC.

Our results align with previous studies that have demonstrated the role of the MAPK pathway in CRC progression and therapy resistance. Oncogenic activation of MAPK signaling, particularly via RAS and BRAF mutations, is known to promote aggressive tumor behavior and confer resistance to EGFR-targeted therapies in CRC patients (20). The observed differences in MAPK pathway alterations between H/L and NHW EOCRC patients may indicate distinct tumor biology, potentially influencing treatment response and survival outcomes. Further research is needed to determine whether H/L patients exhibit differential responses to MAPK-targeted therapies, such as MEK and RAF inhibitors (19, 20).

The survival analysis revealed no significant differences in overall survival for H/L EOCRC patients with or without MAPK pathway alterations (p = 0.9). This suggests that while MAPK mutations are prevalent in this population, their prognostic impact may be limited. In contrast, JAK/STAT pathway alterations in H/L EOCRC patients showed a trend toward worse survival outcomes, though the difference was only borderline significant (p = 0.073). These findings suggest a potential, albeit modest, prognostic role for JAK/STAT pathway disruptions in EOCRC among H/L patients. In NHW EOCRC patients, JAK/STAT alterations were strongly associated with poorer survival outcomes (p < 0.0001), emphasizing their potential prognostic relevance in this ethnic group.

The observed ethnic disparities in MAPK and JAK/STAT pathway alterations may be influenced by genetic ancestry, environmental factors, and lifestyle differences. Studies have shown that genetic ancestry plays a significant role in cancer susceptibility and tumor biology, with H/L individuals exhibiting unique mutational profiles in various cancers, including CRC (6–10, 22, 23). Additionally, dietary and lifestyle factors, such as higher consumption of processed foods and reduced access to preventive healthcare, may contribute to CRC risk and progression in this population (9). Future research integrating genetic, environmental, and clinical data will be essential to elucidate the complex interactions driving CRC disparities.

The implications of our findings for precision medicine are significant. Given the high prevalence of MAPK pathway alterations in H/L EOCRC patients, therapies targeting this pathway, such as MEK inhibitors, may hold promise for this population (16, 17). Additionally, the potential prognostic impact of JAK/STAT pathway alterations justify further investigation into the efficacy of JAK inhibitors in CRC treatment. The lack of significant survival differences associated with MAPK alterations suggests that additional biomarkers, such as tumor microenvironment characteristics and immune signatures, should be explored to refine prognostic models and therapeutic strategies for H/L CRC patients (25, 26).

Despite the strengths of this study, including the use of large-scale genomic data and rigorous statistical analyses, several limitations must be acknowledged. First, the retrospective nature of bioinformatics analyses may introduce selection bias, as publicly available genomic databases may not fully represent the broader H/L CRC population. Second, the relatively small sample size of H/L EOCRC patients may limit the statistical power to detect subtle differences in mutation frequencies and survival outcomes. Third, the lack of functional validation studies prevents direct mechanistic insights into how MAPK and JAK/STAT alterations contribute to CRC disparities.

Future studies should address these limitations by conducting prospective cohort studies with larger, more diverse H/L CRC populations. Additionally, integrating multi-omics approaches, such as transcriptomics and proteomics, will provide a deeper understanding of the functional consequences of MAPK and JAK/STAT pathway disruptions in EOCRC. Investigating potential interactions between these pathways and other molecular drivers, such as WNT and PI3K signaling (7), will be crucial for developing targeted therapies tailored to H/L CRC patients.

In conclusion, this study highlights significant ethnic-specific differences in MAPK pathway alterations in EOCRC, with H/L patients exhibiting a higher prevalence of key mutations compared to NHW patients. While JAK/STAT pathway alterations were not significantly different between ethnic groups, their potential prognostic impact in H/L patients justify further investigation. These findings underscore the importance of precision medicine approaches that account for genetic and ethnic heterogeneity in CRC, with the goal of improving treatment outcomes and reducing cancer disparities in underrepresented populations.

## 5. Conclusions

In conclusion, our findings provide new insights into the ethnicity-specific molecular alterations in the MAPK and JAK/STAT pathways in EOCRC, particularly among H/L patients. The significantly higher prevalence of key MAPK pathway alterations, including NF1, ACVR1, and MAP2K1, in H/L EOCRC patients suggests a potential role for MAPK dysregulation in CRC disparities. Additionally, while JAK/STAT pathway alterations were not significantly different between H/L and NHW patients, their borderline association with survival outcomes in H/L EOCRC patients justify further investigation. These findings underscore the need for further research into the molecular drivers of EOCRC disparities, as well as the potential clinical implications of MAPK and JAK/STAT pathway disruptions. The observed ethnic-specific differences highlight the importance of precision medicine approaches tailored to diverse populations. Future studies should integrate multi-omics analyses and clinical outcomes to refine targeted therapies that address the unique tumor biology of H/L CRC patients. By advancing our understanding of the genetic landscape of EOCRC, these insights may contribute to reducing cancer disparities and improving treatment strategies for underrepresented populations.

## Data Availability

All data used in the present study is publicly available at https://www.cbioportal.org/ and https://genie.cbioportal.org. Additional data can be provided upon reasonable request to the authors.

